# Xpert MTB/RIF Ultra resistant and MTBDR*plus* susceptible rifampicin results in people with tuberculosis: utility of FluoroType MTBDR and deep sequencing

**DOI:** 10.1101/2024.10.25.24316070

**Authors:** Yonas Ghebrekristos, Aysha Ahmed, Natalie Beylis, Sarishna Singh, Christoffel Opperman, Fahd Naufal, Megan Folkerts, David Engelthaler, Erick Auma, Rouxjeane Venter, Ghowa Booley, John Metcalfe, Robin Warren, Grant Theron

## Abstract

**Background:** Xpert MTB/RIF Ultra (Ultra)-detected rifampicin-resistant tuberculosis (TB) is often programmatically confirmed using MTBDR*plus*. There are limited data on discordant results, including re-tested using newer methods like FluoroType MTBDR (FT-MTBDR) and targeted deep sequencing.

**Methods:** MTBDR*plus* rifampicin-susceptible isolates from people with Ultra rifampicin-resistant sputum were identified from a South African programmatic laboratory. FT-MTBDR and single molecule-overlapping reads deep (SMOR; *rpoB, inhA, katG*) on isolate DNA were done (SMOR reference standard).

**Findings:** Between 01/04/2021-30/09/2022, 8% (109/1347) of Ultra rifampicin-resistant specimens were MTBDR*plus*-susceptible. Of 89% (97/109) isolates with a sequenceable *rpoB*, SMOR resolved most in favour of Ultra [79% (77/97)]. Sputum with lower mycobacterial load was associated with Ultra false-positive resistance [46% (11/24) of “very low” Ultras had false-resistance vs. 12% (9/73; p=0.0004) in those ≥“low”], as were Ultra heteroresistance calls (all wild type probes, ≥1 mutant probe) [62% (23/37 vs. 25% (15/60) for Ultra without heteroresistance calls; p=0.0003]. Of the 91% (88/97) of isolates successfully tested by FT-MTBDR, 55% (48/88) were FT-MTBDR rifampicin-resistant and 45% (40/88) susceptible, translating to 69% (47/68) sensitivity and 95% (19/20) specificity. In the 91% (99/109) of isolates with *inhA* and *katG* sequenced, 62% (61/99) were SMOR isoniazid-susceptible.

**Interpretation:** When Ultra and MTBDR*plus* rifampicin results are discordant, Ultra is more likely to be correct and FT-MTBDR agrees more with Ultra than MTBDR*plus*, however, lower load and the Ultra heteroresistance probe pattern were risk factors for Ultra false rifampicin-resistant results. Most people with Ultra-MTBDR*plus* discordant resistance results were isoniazid-susceptible. These data have implications for drug-resistant TB diagnosis.

**Funding:** This work was supported by European & Developing Countries Trial Partnerships (EDCTP2; RIA2020I-3305, CAGE-TB), National Institutes of Health (D43TW010350; U01AI152087; U54EB027049; R01AI136894).

## Introduction

Xpert MTB/RIF Ultra (Ultra; Cepheid, Sunnyvale, USA) is a widely used test for diagnosing tuberculosis (TB) and detecting rifampicin-resistance. Endorsed by the WHO, Ultra has been an essential screening tool in high-incidence countries, such as South Africa, where it has been used routinely since 2011. At the time of this study, in the South African TB Control programme, if Ultra detected rifampicin-resistance, a second specimen is typically cultured. The resulting TB isolate can be tested with GenoType MTBDR*plus* VER 2.0 (MTBDR*plus*, Bruker-Hain Lifescience, Nehren, Germany) which confirms the rifampicin-resistance and can additionally detect isoniazid resistance. However, discrepancies between results obtained directly from the patient specimen and those from cultured isolates or other molecular assays can occur, complicating both reporting and clinical management. This discordance, which could be due to heteroresistance, can lead to poor outcomes,^1^ patient distress and significant financial burdens due to delays and additional testing.

Although superseded in some settings by FluoroType MTBDR VER 2.0 (FT-MTBDR; Bruker-Hain Lifescience, Nehren, Germany),^2, 3^ MTBDR*plus* is widely used for confirmatory drug susceptibility testing (DST). Both Ultra and MTBDR*plus* target the *rpoB* rifampicin-resistant determining region (RRDR) of *Mycobacterium tuberculosis* complex (MTBC). Reports from our high burden setting of South Africa highlight discordance in ~ 7% of Xpert MTB/RIF (Xpert; Ultra’s predecessor) resistant samples, which were MTBDR*plus* rifampicin-susceptible.^4, 5^ Sub-optimal Xpert readouts, particularly in the “very low” semi-quantification category, and probe delay have been linked to false-rifampicin-resistance calls.^6^ However, this has not yet been studied in the context of Ultra. Aside from factors like human error or cross-contamination, discordant results may also arise due to heteroresistance and culture bias, as Ultra is performed directly on specimens while MTBDR*plus* is typically conducted on cultured isolates.^7-9^

Two additional critical gaps exist. First, although Ultra itself does not directly report rifampicin heteroresistance, probe melting temperatures have been suggested as a potential tool for inferring heteroresistance (if a specific probe has melting temperatures corresponding to both wild-type and mutant strains).^10^ However, the diagnostic accuracy of such Ultra’s heteroresistance calls on clinical specimens has not been evaluated. Second, it is unclear whether the level of discordance between Ultra and FT-MTBDR, which utilizes LiquidArray technology to detect MTBC and mutations in *rpo*B, *inh*A, and *kat*G genes, is comparable to that observed with MTBDR*plus*.^2^

We sought to address these knowledge gaps in individuals identified programmatically as having discordant rifampicin results (Ultra-resistant, MTBDR*plus*-susceptible). To ascertain true rifampicin susceptibility status, we employed targeted deep sequencing with single molecule-overlapping reads (SMOR) as a reference standard.^7, 9^ Additionally, we used FT-MTBDR as a comparator. Our study also sought to identify test parameters associated with discordance, heteroresistance, and rifampicin mono-resistance.

## Materials and methods

### Study design and setting

The study was conducted from 1 April 2021 to 30 September 2022, using patient specimens and their corresponding isolates processed at the high-throughput National Health Laboratory Service (NHLS) Greenpoint TB Laboratory (Cape Town, South Africa; ~60 000 TB tests per month).

### Routine diagnostic algorithm

Following the diagnostic algorithm, healthcare workers collected two sputum samples an hour apart from individuals with presumptive TB. Upon laboratory receipt, one specimen was arbitrarily selected for testing with Ultra and processed according to the manufacturer’s instructions.^11^ If Ultra detected MTBC and rifampicin-resistance, the second specimen was processed for mycobacterial culture using the standard NALC-NaOH (1.25% final concentration) decontamination procedure and 0.5 ml inoculated into a Mycobacterium Growth Indicator Tube 960 (MGIT960; Becton Dickinson Diagnostic Systems, Sparks, USA) supplemented with polymyxin B (400 units/ml), amphotericin B (40 µg/ml), nalidixic acid (160 µg/ml), trimethoprim (40 µg/ml) and azlocillin (40 µg/ml) (PANTA, Becton Dickinson Diagnostic Systems) and incubated for ≤35 days. After a tube is automatically flagged growth-positive, Ziehl-Neelsen (ZN) microscopy was performed to detect acid-fast bacilli (AFB). If AFBs were observed, MTBDR*plus* was conducted on the MGIT culture according to the manufacturer’s protocol, using the GenoScan instrument with semi-automated reading and manual confirmation.^12^ All MGIT isolates were stored at room temperature.

### Discordant isolate selection and definition of Ultra heteroresistance results

We selected MTBDR*plus* rifampicin-susceptible isolates from specimens collected concurrently with those tested by Ultra **(Figure 1)**. Patients were classified as Ultra heteroresistant based on the melting temperature curve peaks for each *rpoB* probe as previously described.^10^ Briefly, if each probe exhibited melting peaks corresponding to the wild-type temperature in addition to at least one *rpoB* mutant melting peak, the result was designated heteroresistant (**Figure 2**).

**Figure 1.**
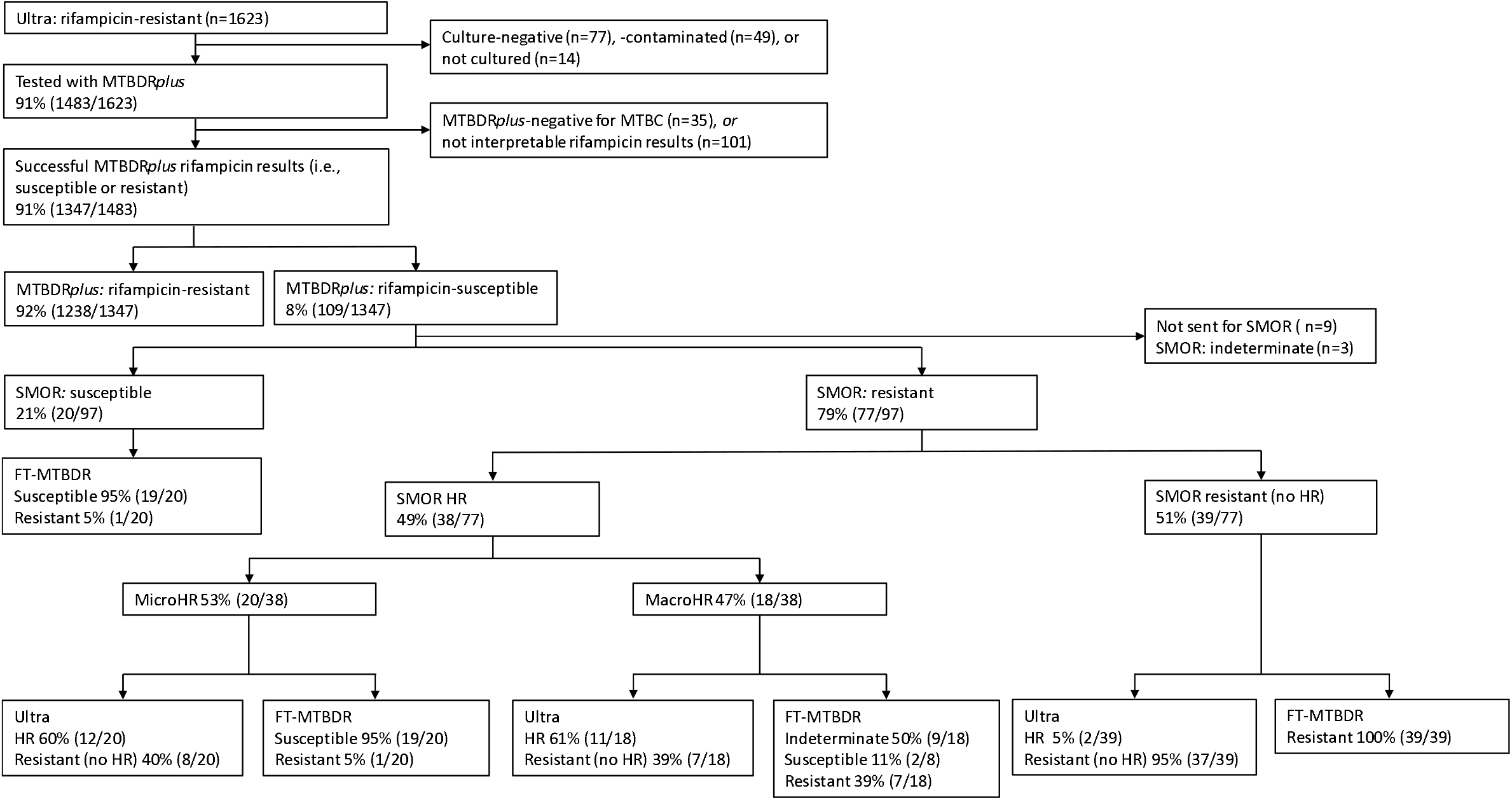
Study profile. We quantified discordant rifampicin susceptibility results (Ultra-resistant, MTBDR*plus*-susceptible) done at TB diagnosis on respiratory specimens over an 18-month period. The distribution of HR is shown, and most isolates that were confirmed by sequencing have RAVs missed by MTBDR*plus* but often detected by FT-MTBDR. Abbreviations: FT-MTBDR, FluoroType MTBDR; MicroHR, microheteroresistance; MacroHR, macroheteroresistance; MTBC, *Mycobacterium tuberculosis* complex; SMOR, single molecule-overlapping repeats; RAV, resistance-associated variant; TB, tuberculosis; Ultra, Xpert MTB/RIF Ultra.

**Figure 2.**
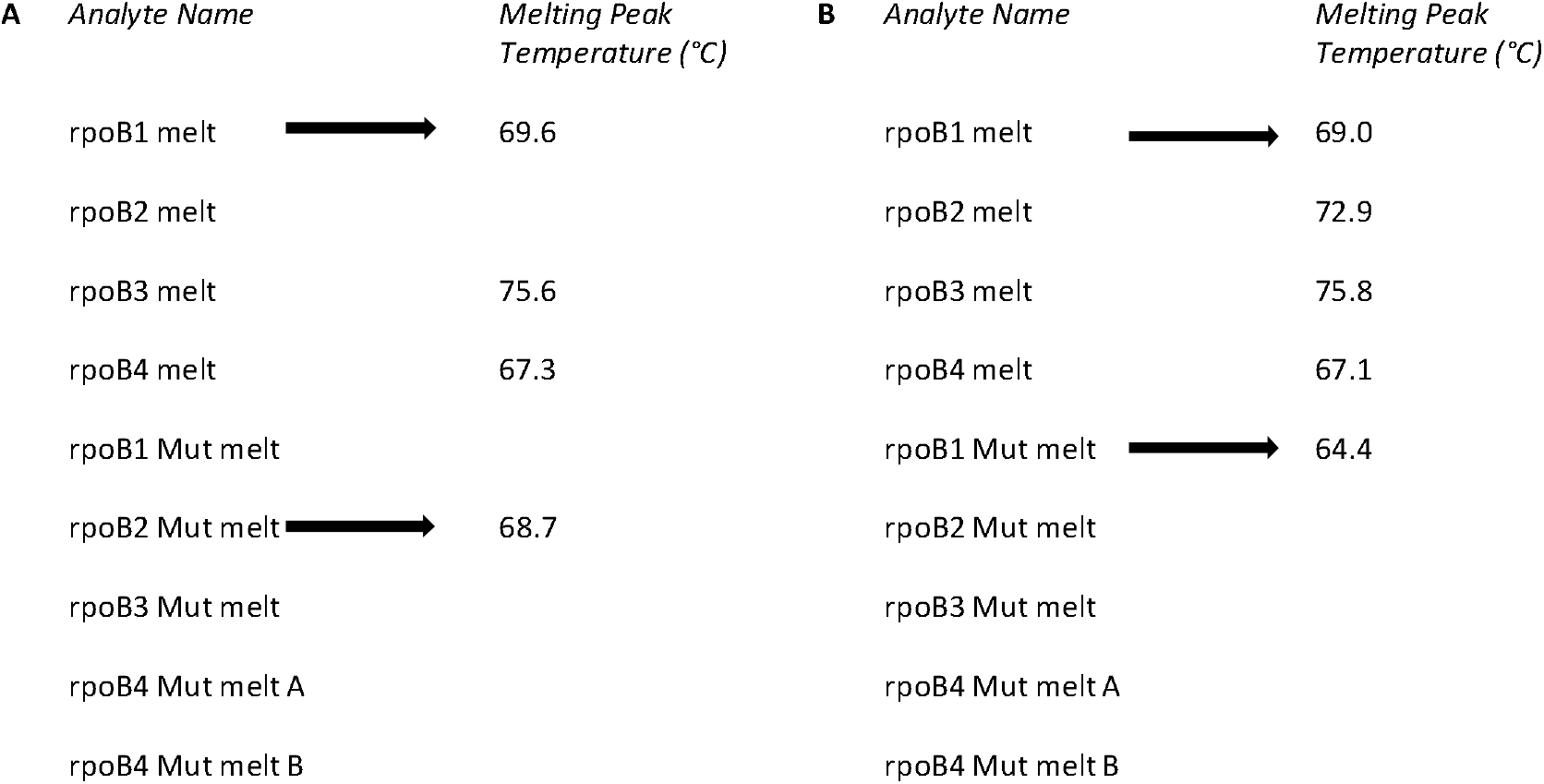
Examples of an Ultra report generated by the GeneXpert software showing melt peak temperatures for each amplicon. **(A)** is a commonly seen rifampicin-resistant specimen with a variant in the rpoB2 region. There is no indication of HR because, for rpoB2, “melt” has no value but “mut melt” does (black arrows). In contrast, **(B)** has, for rpoB1, both “melt” and “mut melt” values, suggesting HR. Abbreviations: °C; degree Celsius; HR, heteroresistance; melt, wild type melt peak temperature; mut melt, mutant melt peak teak temperature; Ultra, Xpert MTB/RIF Ultra.

### FluoroType MTBDR

*DNA Extraction*: 500 µl of MGIT960 growth culture was treated with 167 µl of inactivation reagent (room temperature, 30 min) and DNA was extracted using the GXT96 X2 Extraction Kit VER1.0 (Bruker-Hain Lifescience, Nehren, Germany) with the GenoXtract fleXT instrument (Bruker-Hain Lifescience, Nehren, Germany), as per the manufacturer’s instructions.^13^ With each extraction, saline buffer and un-inoculated MGIT960 (supplemented with PANTA) were included as a negative control, alongside the provided positive control.

*PCR*: Extracted DNA was amplified using the FluoroCycler XT (Bruker-Hain Lifescience, Nehren, Germany) and analysed with the controls using FluoroSoftware XT-IVD (version 1.0.1.5.5.75; Bruker-Hain Lifescience, Nehren, Germany).

### Single Molecule Overlapping Reads (SMOR)

#### DNA extraction

Briefly, 100 μl of growth from the MGIT960 tube was heated at 100°C for 30 min, as previously described.^14^

#### Sequencing

Primers were used to amplify *inhA, katG*, and *rpoB* resistance determining regions as described **(Supplementary Material pp2)**.^15^ A second PCR added adapters using a previously published universal tail method. Samples were pooled and sequenced on an Illumina MiSeq (V3, 600bp paired-end chemistry). Multiple no template controls were used as quality control to ensure the integrity of results.

#### Bioinformatics

The Amplicon Sequencing Analysis Pipeline (version 1.9; ASAP) was used,^15, 16^ which requires overlapping forward and reverse reads to agree and uses read counts to report variant frequency. Resistance calls were classified by ASAP into predefined categories based on the percentage of reads with a known resistance associated variant (RAV) as “microheteroresistance” (0.1-<5%), “macroheteroresistant” (5-95%) or “full resistant” (>95%).^8^ SMOR requires at least 10 paired reads at a locus to make a call. In this case, to call to 0.1%, 10,000 paired reads were required for reporting. When multiple RAVs were detected in a single amplicon, ASAP was used to determine whether they were on the same read as previously described^17^ and thus likely originate from a single population (haplotype identification).

### Statistical analysis and definitions

2×2 tables were used to calculate sensitivity and specificity with 95% confidence intervals (CIs, exact binomial method) and using SMOR results as a reference standard for rifampicin and isoniazid. The prtesti command (STATA 18, StataCorp) was used for comparisons between proportions. Results were classified as successful if a test yielded a definitive resistant or susceptible result; outcomes where MTBC was not detected, or results were uninterpretable were considered unsuccessful.^18^

### Ethics

This study was done according to relevant guidelines and regulations, and approved by the Human Research Ethics Committee, Division of Molecular and Human Genetics, Department of Biomedical Sciences at Stellenbosch University, Cape Town, South Africa (S20/08/189) and the National Health Laboratory Service Academic Affairs, Research and Quality Assurance, South Africa (PR2119347). Permission was granted to access anonymised to-be-discarded residual samples collected as part of routine diagnostic practice with waived informed consent.

## Results

### Frequency of discordant rifampicin results

Between 01/04/2021-30/09/2022, 1623 patients with Ultra rifampicin-resistance were identified. MTBDR*plus* was performed on 91% (1483/1623) of these samples, with 91% (1347/1483) yielding determinate results for rifampicin susceptibility. Of these, 8% (109/1347) were MTBDR*plus* rifampicin-susceptible, and hence discordant with Ultra **(Figure 1)**.

### Relationship between Ultra and SMOR rifampicin results

*RAV frequency:* Of 92% (100/109) Ultra-MTBDR*plus* discordant isolates available for SMOR, 97% (97/100) generated a successful result; 39% (38/97) were classified as full resistant, 19% (18/97) macroheteroresistant, 21% (20/97) microheteroresistant, and the remainder 21% (20/97) had no resistance-associated reads. Therefore, the positive predictive value (PPV) of Ultra rifampicin-resistance for true rifampicin-resistance (as defined by a SMOR reference standard) was 79% (77/97), with 21% (20/97) of Ultra results correspondingly being false-positive for rifampicin-resistance. Lower Ultra-detected load (higher C_Tmin_) was positively associated with false-positive results [median (IQR) C_Tmin_ 29 (28-31) vs. 19 (18-25) in true positives; p=0.0001]. Specifically, in Ultra results with a “very low” semi-quantitation category, 46% (11/24) had false resistance compared to 12% (9/73 p=0.0004) in those with a higher semi-quantitation category [when restricted to those with Ultra heteroresistant patterns these were 75% (6/8) vs. 21% (6/29); p=0.0037] (**Table 1**).

**Table 1.** Ultra parameters among true- and false-Ultra rifampicin-resistant results, in people who were MTBDR*plus*-susceptible, using SMOR on DNA from isolates as a reference standard. Ultra heteroresistance calls and lower detected bacillary load are more frequent among false-resistance results. Data are median (IQR) or % (n/N).

|  | True rifampicin-resistant<br>(n=77) | False rifampicin-resistant<br>(n=20) |
| --- | --- | --- |
| **C<sub>Tmin</sub>** | 19 (18-25) | 29 (27-31); **p=0.0001** |
| ***Semi-quantitation category*** |  |  |
| High | 40 (31/77) | 5 (1/20); p=0.0028 |
| Medium | 31 (24/77) | 15 (3/20); p=0.1506 |
| Low | 12 (9/77) | 25 (5/20); p=0.1312 |
| Very low | 17 (13/77) | 55 (11/20); **p=0.0004** |
| ***Specific probes with mutation label*** |  |  |
| rpoB1 Mut | 47 (32/68) | 41 (7/17); p=0.6633 |
| rpoB2 Mut | 13 (9/68) | 18 (3/17); p=0.6403 |
| rpoB3 Mut | 4 (3/68) | 12 (2/17); p=0.2491 |
| rpoB4 Mut A | 13 (9/68) | 12 (2/17); p=0.8716 |
| rpoB4 Mut B | 29 (2/68) | 6 (1/17); p=0.5567 |
| More than one MUT probe | 19 (13/68) | 12 (2/17); p=0.4769 |
| ***Ultra heteroresistance pattern*** |  |  |
| Heteroresistance pattern | 32 (25/77) | 60 (12/20); **p=0.0239** |

Abbreviations: C<sub>Tmin</sub>, cycle threshold minimum; HR, heteroresistance; MUT, mutation; Ultra, Xpert MTB/RIF Ultra. P-values are for within row comparisons across columns.

### Heteroresistance

Thirty-nine percent (38/97) of people had SMOR-detected heteroresistance and 38% (37/97) of Ultra results exhibited heteroresistant probe patterns. Among these Ultra results, 67% (25/37) had SMOR- detected resistance with two classified as resistant, 11 as macroheteroresistant, and 12 as microheteroresistance; 12 were classified as susceptible by SMOR. Of the 60 Ultra results without heteroresistant patterns, 87% (52/60) had SMOR-detected resistance with 37 classified as resistant, seven as macroheteroresistant, and eight as microheteroresistant; eight were classified as susceptible by SMOR. SMOR-detected heteroresistance was more common in Ultra-detected heteroresistance isolates compared to those without Ultra-detected heteroresistance [62% (23/37) vs. 25% (15/60); (p=0.0003)]. Ultra heteroresistance patterns therefore had 61% (23/38) sensitivity and 95% (37/39) specificity for SMOR heteroresistance. Lastly, Ultra heteroresistance was more likely than Ultra non-heteroresistance resistance results to be false-positive for rifampicin-resistance [PPV, 68% (25/37) vs 87% (52/60); p=0.021].

### Haplotyping

Isolates from 21% (16/77) people had two or more *rpo*B mutations detected by SMOR. Nineteen percent (3/16) had all mutant calls on the same read and the remaining 79% (13/16) had mutations on separate reads, suggesting they were in separate strain subpopulations.

### Isoniazid susceptibility

Thirty-eight of 99 (38%) samples demonstrated isoniazid resistance-associated mutations by SMOR (22 resistant, 7 macroheteroresistant, 9 microheteroresistance; 61 susceptible); 55% (21/38) had *kat*G and 45% (17/38) *inh*A mutations by SMOR. Sensitivity and specificity for isoniazid resistance by MTBDR*plus* were 53% (20/38) and 98% (60/61), respectively **(Supplementary Material Table 2)**. Among the isolates that were false MTBDR*plus* isoniazid-susceptible, 67% (12/18) had heteroresistance (8 microheteroresistance). Heteroresistance was less frequent in MTBDR*plus* isoniazid true positives, with 20% (4/20) being heteroresistant (3 macroheteroresistance). Notably, of the 77 isolates that were SMOR rifampicin-resistant, 56% (43/77) were isoniazid-susceptible (rifampicin mno-resistant).

### FluoroType MTBDR

#### Rifampicin

From the usable rifampicin SMOR results, 91% (88/97) also had successful FT-MTBDR results with 55% (48/88) resistant (Ultra-concordant) and 45% (40/88) susceptible (MTBDR*plus*-concordant). FT-MTBDR sensitivity and specificity for rifampicin-resistance were 69% (47/68) and 95% (19/20), respectively **(Supplementary Material Table 2)**. Amongst FT-MTBDR rifampicin-susceptible isolates, 53% (21/40) were rifampicin-resistant via SMOR. Of these, 100% (21/21) had heteroresistance (19 microheteroresistant) and in all 100% (47/47) SMOR rifampicin-resistant isolates without heteroresistance were detected correctly by FT-MTBDR.

#### Isoniazid

From the 98 people with successful isoniazid FT-MTBDR and SMOR results, 72% (71/98) were FT-MTBDR susceptible and 28% (27/98) FT-MTBDR resistant. FT-MTBDR sensitivity and specificity for isoniazid resistance were 71% (27/38) and 100% (60/60); respectively **(Supplementary Material Table 2)**. Among FT-MTBDR isoniazid-susceptible isolates, 15% (11/71) were isoniazid-resistant by SMOR. All of these, 100% (11/11) had heteroresistance (8 microheteroresistant) and 100% (22/22) SMOR isoniazid-resistant without heteroresistance were detected correctly by FT-MTBDR. Among people with SMOR heteroresistance, FT-MTBDR correctly detected resistance in 31% (5/16).

#### Compared to MTBDRplus for isoniazid resistance

99 people had successful MTBDR*plus* and FT-MTBDR results, 90% of which were concordant (19 resistant, 70 susceptible) and 10 discordant [8 FT-MTBDR resistant and MTBDR*plus* susceptible, 2 FT-MTBDR susceptible and MTBDR*plus* resistance; SMOR supported the FT-MTBDR result in 90% (9/10) people]. The sensitivity of FT-MTBDR for isoniazid resistance was better than MTBDRp*lus* [53% (20/38) vs 71% (27/38); p=0.0983], whereas specificity remained similar [98% (60/61) vs 100% (60/60); p=0.3193] **(Supplementary Material Table 2)**.

## Discussion

To our knowledge, this study is the first to describe rifampicin susceptibility discordance between the WHO-recommended rapid molecular tests Ultra and MTBDR*plus*. Our key findings are: 1) Most discordance (79%) was from MTBDR*plus* not detecting rifampicin-resistance and 2) 69% of these MTBDR*plus*-susceptible were detected as FT-MTBDR resistant, indicating FT-MTBDR has higher sensitivity than MTBDR*plus*. However, 3) a substantial proportion with sequencing-detected resistance (31%; all of which were heteroresistant) were missed by FT-MTBDR and, in people with heteroresistance, multiple resistant strains were often present. Furthermore, 4) although the Ultra heteroresistance probe pattern was associated with heteroresistance, this pattern had suboptimal sensitivity and specificity for heteroresistance and was itself associated with Ultra false-resistant calls (as was lower mycobacterial load). Lastly, 5) more than half of the individuals were rifampicin mono-resistant, supporting the need for isoniazid DST. These data have implications for laboratory DST algorithms, especially resolution of discordant results by different molecular methods.

Most rifampicin resistance discordance arose from MTBDR*plus* not detecting RAVs, rather than Ultra falsely detecting RAVs. This might be because MTBDR*plus* interpretation is subjective even with the semi-automated GenoScan and requires human reporting. In contrast, FT-MTBDR reporting is fully automated. While FT-MTBDR identified most resistance missed by MTBDR*plus*, approximately half of the isolates FT-MTBDR detected as rifampicin-susceptible had sequencing-detected resistance. This contrasts with other studies that have reported FT-MTBDR sensitivities approaching 100%,^2^ however, these were done in Ultra rifampicin-resistant people (without specifically selecting the discordant MTBDR*plus*-susceptible subset).

Heteroresistance, which we show to be a cause of Ultra-MTBDR*plus* discordance was, about a third of the time, missed by FT-MTBDR. However, as these people were MTBDR*plus*-susceptible, FT-MTBDR is still substantially better at detecting resistance than the previous generation technology. Interestingly, within people with sequencing-detected heteroresistance, there was seldom one resistant strain implicated, which is unexpected given that these are not samples taken from people on treatment and sequencing was done after culture, which can result in loss of minority variants.^8^ Possible causes of this diversity include multiple exposures to rifampicin-resistant TB or substantial intra-host evolution.

Certain probe patterns reported by Ultra have been proposed to be useful for diagnosing heteroresistance,^10^ which may be clinically useful if first-line drugs could be included in the regimen to rapidly reduce bacterial load of the drug-susceptible subpopulation.^19, 20^ However, in our study, although this Ultra probe pattern was indeed associated with heteroresistance, it did not translate into high sensitivity and specificity for heteroresistance. While FT-MTBDR does not currently offer a heteroresistance readout, this feature could be incorporated into its software to potentially inform treatment. Lastly, this Ultra hetereoresistance pattern (as well as that from FT-MTBDR) was also associated with Ultra false-resistance calls, as was the Ultra “very low” semi-quantitation category. This category is a recognized risk factor for Xpert false-resistance,^6^ for which repeat testing is recommended. Our data therefore suggests that samples with an Ultra heteroresistance pattern and or “very low” bacterial load should be considered at increased risk for false-resistance. The utility of repeat testing in such samples warrants further evaluation.

Our findings emphasize the importance of not assuming rifampicin-resistance equates to isoniazid resistance, particularly in cases of discordant Ultra-MTBDR*plus* results. Previous studies have demonstrated 19%^21^ and 21%^22^ of Xpert rifampicin-resistant cases are isoniazid-susceptible by MTBDR*plus*. Our data therefore support the scale-up of upfront routine isoniazid DST to avoid the inappropriate exclusion of isoniazid from regimens.

Our study has strengths and limitations. Ultra was done on specimens, while MTBDR*plus*, FT-MTBDR and SMOR were done on isolates. While this may be representative of some programmatic algorithms, changes in subpopulation structures due to culture bias could create discordance. Furthermore, phenotypic susceptibility testing was not possible as isolates were not stored to preserve viability. However, SMOR has high sensitivity and specificity for phenotypic (and sub-phenotypic) resistance.^7^ Another consideration is that our study was designed to investigate Ultra-resistant, MTBDR*plus-*susceptible discordance, rather than to assess Ultra rifampicin resistant calls in all comers (which others have done for Ultra’s predecessor Xpert).^6^ In other words, our findings should be interpreted within the context of samples pre-selected because they were Ultra-MTBDR*plus* discordant (such discordant samples are likely not representative of typical RAVs in our setting).

In conclusion, patients with Ultra rifampicin-resistant TB that were susceptible by MTBDR*plus* are predominantly truly rifampicin resistant unless Ultra produced a heteroresistant probe pattern or “very low” semi-quantitation category (both associated with false Ultra rifampicin results). Isoniazid, for which susceptibility testing should be done, likely remains useful in people with this Ultra-MTBDR*plus* discordance.

## Data Availability

Study data can be accessed on reasonable request from the corresponding author without restriction.

## Author Contributions

All named authors were involved in conceptualization and design of the study. Y.G., A.A., G.B. and E.A. assisted with specimen collection and specimen processing. F.N. M.F. and D.E. performed SMOR testing. Y.G., G.T., E.A., R.V., M.F., F.N. and D.E. performed formal analysis, methodology and writing original draft. All authors had full access to the de-identified data. All authors reviewed and edited the manuscript and had final responsibility for the decision to submit for publication.

## Data Sharing

Study data can be accessed on request from the corresponding author without restriction.

## Acknowledgments

The authors thank the National Health Laboratory (NHLS) as a source of data and the staff of Greenpoint Tuberculosis Laboratory, NHLS, Cape Town, South Africa for assisting with collecting samples. We would like to thank the staff from Clinical Mycobacteriology and Epidemiology (CLIME) Group, Division of Molecular and Human Genetics, Stellenbosch University for their support in performing FT-MTBDR. Hain-Bruker Lifesciences donated FT-MTBDR consumables and loaned the GenoXtract fleXT and FluoroCyclerXT-96 equipment but had no other role in the study. T-Gen provided instrumentation and reagents needed for SMOR processing.

## References

1. Shin SS, Modongo C, Baik Y, Allender C, Lemmer D, Colman RE, et al. Mixed Mycobacterium tuberculosis-Strain Infections Are Associated With Poor Treatment Outcomes Among Patients With Newly Diagnosed Tuberculosis, Independent of Pretreatment Heteroresistance. J Infect Dis. 2018;218(12):1974–82.

2. Dippenaar A, Derendinger B, Dolby T, Beylis N, van Helden PD, Theron G, et al. Diagnostic accuracy of the FluoroType MTB and MTBDR VER 2.0 assays for the centralized high-throughput detection of Mycobacterium tuberculosis complex DNA and isoniazid and rifampicin resistance. Clin Microbiol Infect. 2021;27(9):1351 e1–e4.

3. de Vos M, Derendinger B, Dolby T, Simpson J, van Helden PD, Rice JE, et al. Diagnostic Accuracy and Utility of FluoroType MTBDR, a New Molecular Assay for Multidrug-Resistant Tuberculosis. J Clin Microbiol. 2018;56(9).

4. Ghebrekristos Y. Characterization of Mycobacterium tuberculosis isolates with discordant rifampicin susceptibility test results: University of Cape Town; 2018.

5. Beylis N, Ghebrekristos Y, Nicol M. Management of false-positive rifampicin resistant Xpert MTB/RIF. Lancet Microbe. 2020;1(6):e238.

6. Ngabonziza JCS, Decroo T, Migambi P, Habimana YM, Van Deun A, Meehan CJ, et al. Prevalence and drivers of false-positive rifampicin-resistant Xpert MTB/RIF results: a prospective observational study in Rwanda. Lancet Microbe. 2020;1(2):e74–e83.

7. Metcalfe JZ, Streicher E, Theron G, Colman RE, Allender C, Lemmer D, et al. Cryptic Microheteroresistance Explains Mycobacterium tuberculosis Phenotypic Resistance. Am J Respir Crit Care Med. 2017;196(9):1191–201.

8. Metcalfe JZ, Streicher E, Theron G, Colman RE, Penaloza R, Allender C, et al. Mycobacterium tuberculosis Subculture Results in Loss of Potentially Clinically Relevant Heteroresistance. Antimicrob Agents Chemother. 2017;61(11).

9. Engelthaler DM, Streicher EM, Kelley EJ, Allender CJ, Wiggins K, Jimenez D, et al. Minority Mycobacterium tuberculosis Genotypic Populations as an Indicator of Subsequent Phenotypic Resistance. Am J Respir Cell Mol Biol. 2019;61(6):789–91.

10. Chakravorty S, Simmons AM, Rowneki M, Parmar H, Cao Y, Ryan J, et al. The New Xpert MTB/RIF Ultra: Improving Detection of Mycobacterium tuberculosis and Resistance to Rifampin in an Assay Suitable for Point-of-Care Testing. mBio. 2017;8(4).

11. Xpert® MTB/RIF Ultra (package insert) [Internet]. 2019 [cited 28 February 2023]. Available from: https://www.biovendor.sk/download/30352/Manual_xpert_mtb-rif_ultra.pdf.

12. GenoType MTBDRplus VER 2.0 [Internet].2015 [cited February 2023]. Available from: https://www.hain-lifescience.de/include_datei/kundenmodule/packungsbeilage/download.php?id=936.

13. GXT96 X2 Extraction Kit Version 1 (Package insert) [Internet]. 2022 [cited 2024-07-25]. Available from: https://www.hain-lifescience.de/en/instructions-for-use.html.

14. Kocagoz T, Yilmaz E, Ozkara S, Kocagoz S, Hayran M, Sachedeva M, et al. Detection of Mycobacterium tuberculosis in sputum samples by polymerase chain reaction using a simplified procedure. J Clin Microbiol. 1993;31(6):1435–8.

15. Colman RE, Schupp JM, Hicks ND, Smith DE, Buchhagen JL, Valafar F, et al. Detection of Low-Level Mixed-Population Drug Resistance in Mycobacterium tuberculosis Using High Fidelity Amplicon Sequencing. PLoS One. 2015;10(5):e0126626.

16. Colman RE, Anderson J, Lemmer D, Lehmkuhl E, Georghiou SB, Heaton H, et al. Rapid Drug Susceptibility Testing of Drug-Resistant Mycobacterium tuberculosis Isolates Directly from Clinical Samples by Use of Amplicon Sequencing: a Proof-of-Concept Study. J Clin Microbiol. 2016;54(8):2058–67.

17. Derendinger B, Dippenaar A, de Vos M, Huo S, Alberts R, Tadokera R, et al. Bedaquiline resistance in patients with drug-resistant tuberculosis in Cape Town, South Africa: a retrospective longitudinal cohort study. Lancet Microbe. 2023;4(12):e972–e82.

18. FluoroType MTBDR VER 2.0 (Package insert) [Internet]. Hain Lifescience (Brucker). 2024 [cited 2024-07-25]. Available from: https://www.hain-lifescience.de/en/instructions-for-use.html.

19. Department of Health (DOH) RoSA. Management of rifampicin resistant tuberculosis: A Clinical reference guide. South Africa; 2020.

20. Health WCDo. Clinical Guidelines & Standard Operating Procedure for the Implementation of the Short & Long DR-TB regimens for Adults, Adolescents and Children. South Africa; 2018 September 2018.

21. Bisimwa BC, Nachega JB, Warren RM, Theron G, Metcalfe JZ, Shah M, et al. Xpert Mycobacterium tuberculosis/Rifampicin-Detected Rifampicin Resistance is a Suboptimal Surrogate for Multidrug-resistant Tuberculosis in Eastern Democratic Republic of the Congo: Diagnostic and Clinical Implications. Clin Infect Dis. 2021;73(2):e362–e70.

22. Pillay S, de Vos M, Derendinger B, Streicher EM, Dolby T, Scott LA, et al. Non-actionable Results, Accuracy, and Effect of First- and Second-line Line Probe Assays for Diagnosing Drug-Resistant Tuberculosis, Including on Smear-Negative Specimens, in a High-Volume Laboratory. Clin Infect Dis. 2023;76(3):e920–e9.

