## Supplemental Table for "Xpert MTB/RIF Ultra resistant and MTBDR*plus* susceptible rifampicin results in people with tuberculosis: utility of FluoroType MTBDR and deep sequencing": Supplementary Material.docx

**Supplementary Table of Contents:**

Supplementary Table 1. *Mycobacterium tuberculosis* complex specific primer with universal tail sequences……………………………………………………………………………………………….2

Supplementary Table 2. Sensitivity and specificity of FT-MTBDR and MTBDR*plus* for rifampicin or isoniazid resistance..……………………………………………………………………….……………3

**Table 1.** *Mycobacterium tuberculosis* complex specific primer with universal tail sequences. All oligos are with standard de-salting. The universal tail sequences are highlighted in red and bold with the forward primer sequence differing from the reverse primer sequence.

| **Forward Primer** | **Sequence** (5`-3`) |
| --- | --- |
| rpoBv2fUT1 | **ACCCAACTGAATGGAGC**CGATCACACCGCAGACGTT |
| katGv2fUT1 | **ACCCAACTGAATGGAGC**CCATGAACGACGTCGAAACAG |
| inhAv2fUT1 | **ACCCAACTGAATGGAGC**CCTCGCTGCCCAGAAAGG |
| **Reverse Primer** |  |
| rpoBv2rUT2 | **ACGCACTTGACTTGTCTTC**GTTTCGATCGGGCACATCC |
| katGv2rUT2 | **ACGCACTTGACTTGTCTTC**GCTCTTCGTCAGCTCCCACTC |
| inhAv2rUT2 | **ACGCACTTGACTTGTCTTC**GTCACATTCGACGCCAAACAG |

**Table 2.** Sensitivity and specificity of FT-MTBDR and MTBDR*plus* for rifampicin or isoniazid resistance detection when done on isolates from Ultra rifampicin-resistant MTBDR*plus* rifampicin-susceptible people using SMOR on DNA from isolates as a reference standard. FT-MTBDR sensitivity for each drug exceeded that of MTBDR*plus*. MTBDR*plus* specificity was higher for rifampicin and was lower for isoniazid than that of FT-MTBDR. Data are % (95% CI; n/N).

|  | Sensitivity | Specificity |
| --- | --- | --- |
| FT-MTBDR: Rifampicin | 69 (57-80; 47/68) | 95 (75-100; 19/20) |
| FT-MTBDR: Isoniazid | 71 (54-85; 27/38) | 100 (94-100; 60/60) |
| MTBDR*plus*: Rifampicin | Non-applicable | 100 (83-100; 20/20) |
| MTBDR*plus*: Isoniazid | 53 (39-69; 20/38) | 98 (91-100; 60/61) |
